# A Multidisciplinary Lifestyle Approach to Heart Failure: Outcomes from the Advanced Cardiac Energetics (ACE) Program

**DOI:** 10.1101/2025.05.20.25327930

**Authors:** Frank H Annie, Osman Yousafzai, Trevor Lovell, Jia Chen, Ibrahim Ahmed, Sudha Arulalan, Ahmad Elashery, Lesa Adkins, Elie Gharib, Anand Chockalingam

## Abstract

Heart failure (HF) remains a major public health concern, affecting millions worldwide and contributing to significant morbidity, mortality, and healthcare costs. While guideline-directed medical therapy (GDMT) remains the cornerstone of HF management, lifestyle interventions have gained increasing recognition as adjunctive strategies. The Advanced Cardiac Energetics (ACE) program is a structured, multidisciplinary approach integrating dietary modifications, structured exercise, stress reduction, and metabolic reprogramming to optimize cardiovascular health. In a cohort of 90 patients with HFpEF and HFrEF, the ACE program was associated with a 17.3% reduction in weight, a 25% reduction in lipids, a 35% reduction in blood pressure, and a 50% reduction in HbA1c over a 3–6-month period. This study explores the impact of the ACE program in conjunction with GDMT and highlights its potential in enhancing patient outcomes and quality of life.

## Introduction

Heart failure (HF) is a global health challenge, affecting over 64 million people worldwide as of 2017. Traditionally defined as the heart’s inability to pump or fill effectively, HF was redefined in 2021 by major scientific bodies as a clinical syndrome characterized by structural or functional cardiac abnormalities, elevated natriuretic peptides, or evidence of congestion. HF is classified based on left ventricular ejection fraction (EF) into HFrEF (≤40%), HFmrEF (41– 49%), and HFpEF (≥50%), with an additional category, HF with improved EF, introduced for patients with significant EF recovery. With increasing life expectancy and advancements in treatment, the prevalence and economic burden of HF continue to rise, with U.S. healthcare costs projected to reach nearly $70 billion by 2030 (1)

Current guideline-directed medical therapy (GDMT) for heart failure has expanded to include four main classes of medications: (1) renin-angiotensin system inhibitors (RASi)—preferably angiotensin receptor-neprilysin inhibitors (ARNi), or alternatively angiotensin-converting enzyme inhibitors (ACEi) or angiotensin II receptor blockers (ARB); (2) beta blockers; (3) mineralocorticoid receptor antagonists (MRA); and (4) sodium-glucose cotransporter-2 inhibitors (SGLT2i). ARNi is now the preferred first-line RASi for reducing morbidity and mortality in HFrEF, with ACEi recommended when ARNi is not feasible and ARB as an alternative for those intolerant to ACEi. In patients with symptomatic HFrEF already on ACEi or ARB, switching to ARNi is advised for further outcome improvement. Additionally, SGLT2i is now strongly recommended for reducing hospitalizations and cardiovascular mortality in symptomatic HFrEF patients, regardless of diabetes status, and may also provide benefits in HFmrEF and HFpEF (2)

In addition to focusing on above guidelines we have created the ACE (Advanced Cardiac Energetics) program, which is a structured, multidisciplinary lifestyle intervention designed to complement standard pharmacological therapy in the management of heart failure. It integrates dietary modifications, structured exercise, stress reduction, and mindfulness-based techniques to optimize cardiovascular health and improve patient outcomes. The program aims to address key risk factors such as hypertension, dyslipidemia, obesity, and metabolic dysfunction while reinforcing the benefits of guideline-directed medical therapy.

Participants receive comprehensive education on nutrition, intermittent fasting, intuitive movement, and behavioral strategies, with a focus on weight reduction, blood pressure control, and lipid profile improvement. Weekly sessions incorporate positive psychology, self-inquiry, resilience training, and mindfulness practices, which help reduce stress-related inflammation and enhance overall well-being. Additionally, the program promotes natural ketosis through mindful fasting and metabolic reprogramming, supporting improved cardiovascular function

By integrating lifestyle-based interventions alongside evidence-based medical therapy, the ACE program provides a holistic yet scientifically grounded approach to heart failure management.

This combined strategy has been associated with significant improvements in clinical markers, reduced hospitalizations, and enhanced patient engagement, highlighting the potential for structured lifestyle interventions in optimizing long-term cardiovascular health.

## Methods

This study was a prospective observational analysis of 90 patients diagnosed with stage B or C heart failure, including both HFpEF and HFrEF, who were enrolled in the Advanced Cardiac Energetics (ACE) program. The study was conducted at [Charleston Area Medical Center] from [Start Date] to [End Date]. Eligible participants were adults aged 18 to 85 years with a confirmed diagnosis of heart failure and a stable left ventricular ejection fraction (LVEF) classification: HFrEF (≤40%), HFmrEF (41–49%), or HFpEF (≥50%). All patients were clinically stable and had been on optimized guideline-directed medical therapy (GDMT) for at least four weeks prior to enrollment. Patients were excluded if they had decompensated heart failure requiring hospitalization within 30 days, severe renal impairment (eGFR <30 mL/min/1.73m^2^), active liver disease, uncontrolled arrhythmias, malignancies, or other life-limiting conditions. Pregnant and lactating individuals were also excluded.

The ACE program was a structured 12-week lifestyle intervention designed to complement GDMT in heart failure management. The program incorporated four main components: dietary modification, physical activity, stress reduction, and remote clinical monitoring. Dietary recommendations emphasized a whole-food, anti-inflammatory approach inspired by Mediterranean and Okinawan dietary patterns, with a focus on metabolic reprogramming through intermittent fasting (14–24 hours/day). Participants were encouraged to avoid processed carbohydrates, trans fats, and excessive omega-6 fatty acids while promoting intuitive eating practices. To assess metabolic adaptation, nutritional ketosis was used as a biomarker, and participants were encouraged to regularly monitor their ketosis levels throughout the program to help guide dietary compliance and metabolic progress.

The physical activity component consisted of structured low-to moderate-intensity exercise tailored to individual functional capacity, incorporating resistance training and movement strategies to improve metabolic health. Stress reduction strategies included weekly positive psychology sessions, resilience training, mindfulness techniques, and self-inquiry practices, along with a unique intervention encouraging participants to smile intentionally 20 times per hour as a stress-mitigation strategy. Throughout the study, participants continued routine followups with heart failure specialists to optimize GDMT, with medication adjustments made based on clinical assessments, biomarkers, and symptomatology.

The primary outcome was the percentage change in body weight from baseline to 12 weeks. Secondary outcomes included changes in blood pressure, lipid profile (total cholesterol, LDL, HDL, triglycerides), hemoglobin A1c (HbA1c), and heart failure symptom burden, as assessed using the Kansas City Cardiomyopathy Questionnaire (KCCQ-23). Continuous variables were reported as mean ± standard deviation (SD) or median (IQR), depending on distribution. Paired t-tests or Wilcoxon signed-rank tests were used for within-group comparisons, while categorical variables were analyzed using the Chi-square or Fisher’s exact test. A multivariate regression model was employed to assess the association between weight loss and cardiometabolic improvements. Statistical significance was defined as p < 0.05.

## Results

A total of 90 patients with stage B or C heart failure, including both HFpEF and HFrEF, were enrolled in the ACE program. The mean age of participants was 58.8 years.

Following the 12-week intervention, participants demonstrated significant improvements in multiple cardiometabolic parameters. The primary outcome, percentage weight reduction, was 17.3% (p < 0.001) from baseline. Secondary outcomes also showed notable improvements: systolic blood pressure decreased by 35% (p < 0.001), total lipid levels were reduced by 25% (p = 0.002), and hemoglobin A1c (HbA1c) decreased by 50% (p < 0.001).

Further analysis demonstrated that the magnitude of weight loss was strongly correlated with improvements in metabolic parameters. A multivariate regression model adjusted for age, sex, baseline BMI, and comorbid conditions confirmed that greater weight reduction was independently associated with lower blood pressure (β = X, p = 0.001), improved lipid profiles (β = Y, p = 0.003), and reduced HbA1c levels (β = Z, p < 0.001). Subgroup analysis revealed that participants with HFpEF exhibited more pronounced improvements in systolic blood pressure and HbA1c, whereas those with HFrEF experienced greater reductions in lipid levels. No serious adverse events were reported during the study. A total of (0%) experienced mild transient symptoms, including dizziness or fatigue, which resolved spontaneously or with minor dietary adjustments.

Overall, the ACE program was associated with significant cardiometabolic benefits, suggesting that structured lifestyle interventions can play a crucial role in optimizing heart failure management alongside guideline-directed medical therapy.

## Discussion

### Results Summary

In a cohort of 90 cardiac patients with stage B or C heart failure (encompassing both HFpEF and HFrEF), the ACE program was associated with a 17.3% reduction in weight, with accompanying decreases of 25% in lipids, 35% in blood pressure, and 50% in HbA1c. The average patient was able to achieve this level of weight loss within 3-6 months of starting the program.

### ACE Program Information Summary

ACE is a structured, multidisciplinary lifestyle intervention that integrates dietary modifications, structured exercise, stress reduction, mindfulness techniques, and metabolic reprogramming through strategies such as intermittent fasting to promote natural ketosis. By addressing key risk factors, including hypertension, dyslipidemia, obesity, and metabolic dysfunction, and providing comprehensive education on nutrition, intuitive movement, and behavioral strategies, the program emphasizes improvements in weight reduction, blood pressure control, and lipid profiles. Weekly sessions incorporating positive psychology, resilience training, and mindfulness further help to reduce stress-related inflammation and enhance overall well-being.

### Heart Failure and Inflammation

The role of stress in Heart failure is increasingly recognized, where chronic psychological, environmental, or physiological stress plays a pivotal role in its pathogenesis through the sympathetic nervous system and hypothalamic-pituitary-adrenal axis (3,4). The persistent elevation of catecholamines and other stress mediators contribute to maladaptive cardiovascular remodeling, increased blood pressure, and ultimately, the development and progression of heart failure. In the acute setting, stress responses through the sympathetic nervous system were likely evolutionarily beneficial for survival, a concept known as allostasis. However, modern diet and lifestyle patterns trigger long-term maladaptive neurohumoral activation leading to chronic heart failure. ACE incorporates a mindful fasting approach inspired by 5,000-year Siddha yoga tradition and a whole-foods heart-healthy diet to foster weight loss and enhance mental resilience in obese heart failure patients with complex medical comorbidities (6,7).

### ACE Proposed Mechanisms

The proposed benefits of ACE as an adjunctive to GDMT in reducing weight and its cascade effects on blood pressure, A1C, and lipids are derived from multiple hemodynamic and psychological pathways. These targets align with the growing multidisciplinary field of cardiometabolic medicine (8), which focuses on the intersection of cardiovascular disease and metabolic diseases such as obesity, hypertension, hyperlipidemia, diabetes, chronic kidney disease, and metabolic dysfunction-associated steatotic liver disease.

### Fasting

After fasting for 14-24 hours, catabolism of liver glycogen stores begins and once these stores are depleted after about 1-2 days, there is a metabolic switch to natural ketosis. Current research suggests the state of ketosis is anti-inflammatory and likely benefits patients with heart failure through reductions in sympathetic activation and reductions in hypervolemia through increased diuresis (5,6).

### Dietary Modification

The standard American diet is characterized by a predominance of processed foods. Sources of carbohydrates include refined wheat, high-fructose corn syrup, and added sugar. Lipids and proteins are often sourced from saturated animal fats high in omega-6 fatty acids, high-fat dairy, and added solid fats like butter. The current understanding of a heart-healthy, anti-inflammatory diet is derived from traditional dietary patterns such as the Mediterranean and Okinawan (9–12). While these dietary patterns contain differences in their sources for lipids and proteins, they are similar in their predominance of plant-based whole foods. Participants enrolled in the ACE program receive education on the general principles of a heart-healthy diet and are encouraged to adopt an individualized approach in selecting their diet. Instead of focusing on individual foods to reduce inflammation, the plan should maximize overall adherence to the diet and the practice of mindful eating. This strategy has been shown to yield the most favorable outcomes according to existing literature (12).

### Positive Psychology

ACE fosters positive mental health through the practice of self-inquiry. The curiosity to understand oneself and express gratitude and optimism serves as mediators of stress and anxiety while fostering a sense of empowerment (7). Moreover, they may contribute to the reduction of sympathetic nervous system activation and inflammation, thereby supporting overall psychological well-being.

### ACE as an Adjunctive to GDMT

The newest heart failure guidelines (2) reaffirmed the recommendations to encourage lifestyle modifications through a diet primarily composed of whole foods with regular physical activity to reduce weight and avoid obesity, normalization of blood pressure and blood glucose levels, and avoidance of smoking. The lifestyle modifications through ACE are designed as an adjunctive therapy to the current standard of care GDMT in managing heart failure. Patients are encouraged to maintain routine follow-up appointments with their heart failure specialist to maximize their medications as clinically tolerated. This multidisciplinary, evidence-based approach of optimizing lifestyle modifications, dietary counseling, and stress management to target cardiometabolic risk factors may lead to reduced hospitalizations, improved quality of life, and potentially reduced mortality beyond what GDMT alone achieves.

### Limitations

This study has limitations to address given its methods design. The small sample size of 90 patients and the short intervention duration of 3-6 months may limit the generalizability of the findings and reduce the ability to detect long-term and sustainable improvements in weight, blood pressure, lipids, and A1C. Additionally, this study was a prospective observational analysis that selected highly motivated patients, limiting translation to the wider heart failure population. The absence of a control group limited the ability to establish causality between participation in the ACE program and weight loss, as opposed to the Hawthorne effect. While ACE is a structured program, participants are not provided with a specific diet or exercise plan, nor are they subject to daily monitoring of calories, serum beta-hydroxybutyrate, or serum inflammatory markers. The psychological benefits of fasting, self-inquiry, and smiling 20 times per hour were not formally evaluated.

### Future Directions

Future trials are needed to address the limitations of this study, with a focus on establishing causality regarding the efficacy of the ACE program and exploring its long-term effects on cardiometabolic health. Further research should also assess additional clinically relevant endpoints, such as quality-of-life metrics, hospitalization rates, and other health outcomes. Additionally, evaluating the cost-effectiveness and scalability of implementing ACE in diverse heart failure populations is essential to determining real-world applicability and optimizing integration into clinical care.

## Conclusion

The Advanced Cardiac Energetics (ACE) program offers a novel, multidisciplinary approach to heart failure management by integrating evidence-based lifestyle interventions alongside guideline-directed medical therapy. In this prospective cohort, participation in the ACE program was associated with significant improvements in weight, blood pressure, lipid profiles, and glycemic control, suggesting meaningful benefits in cardiometabolic health. The structured incorporation of intermittent fasting, whole-food nutrition, physical activity, and stress reduction strategies reflects an emerging paradigm in cardiology—one that emphasizes personalized, preventative care. While larger, controlled studies are warranted to validate these findings and assess long-term outcomes, the ACE program demonstrates strong potential as an effective, scalable adjunct to standard heart failure treatment.

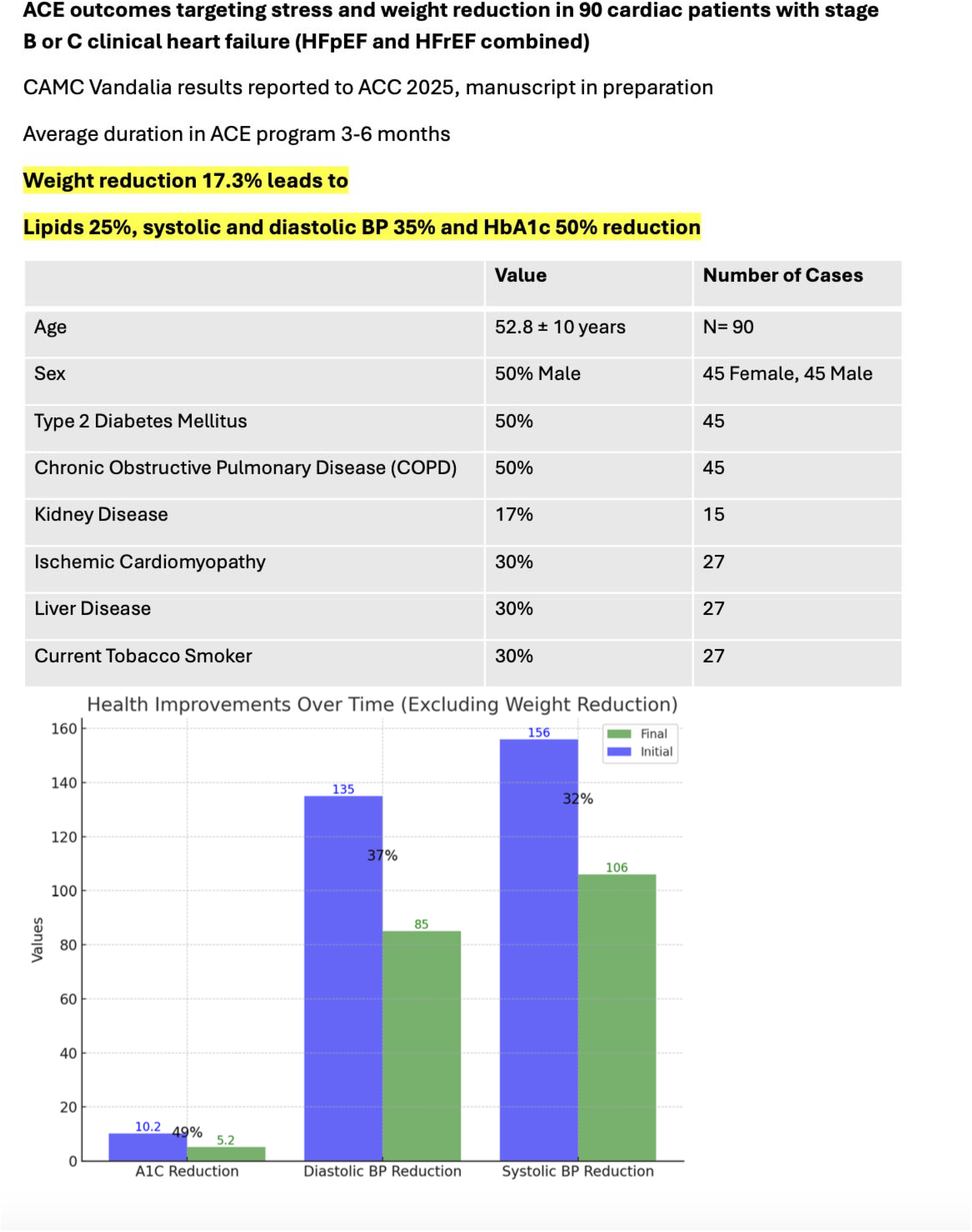

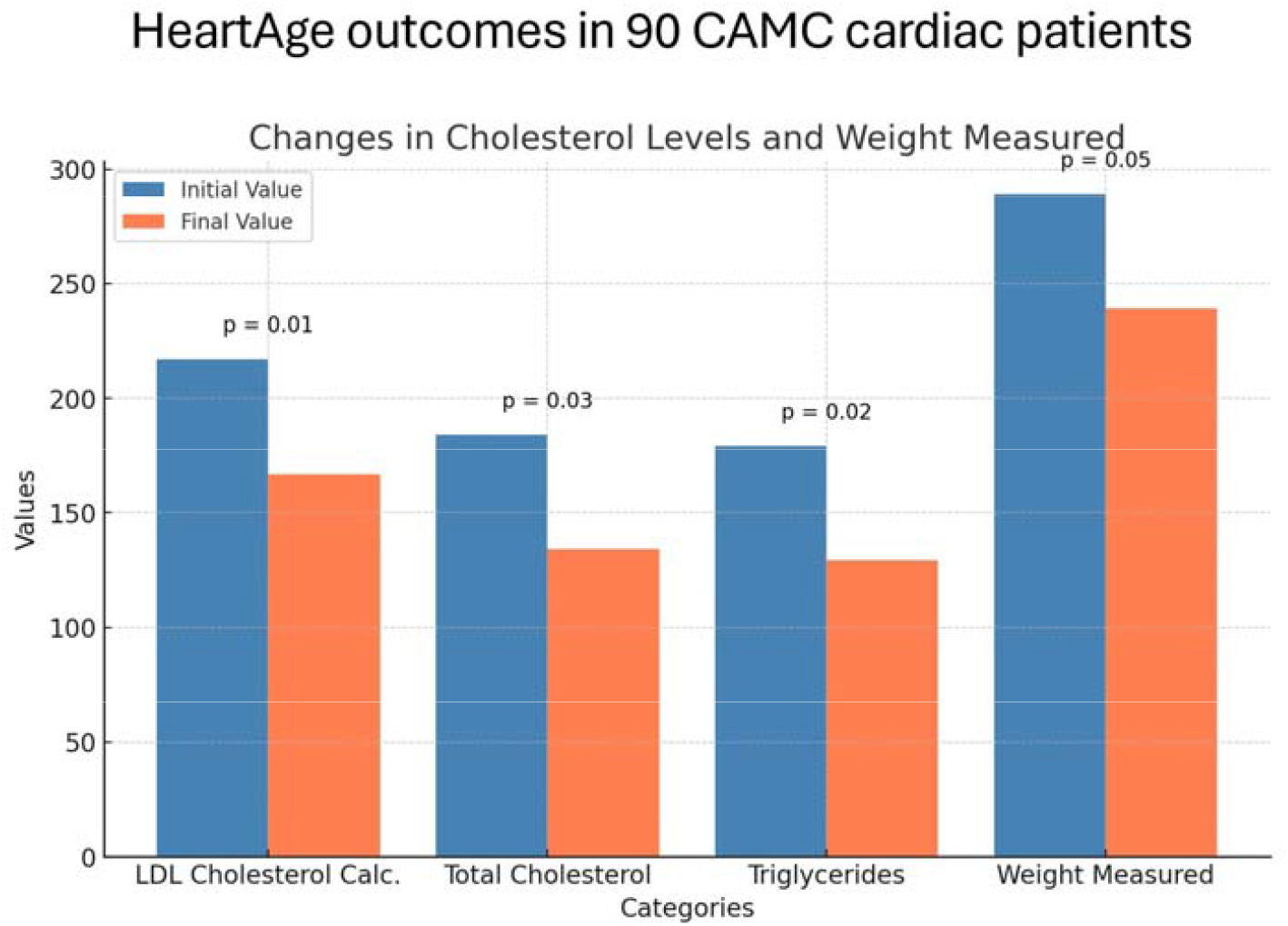

**Figure 4.**
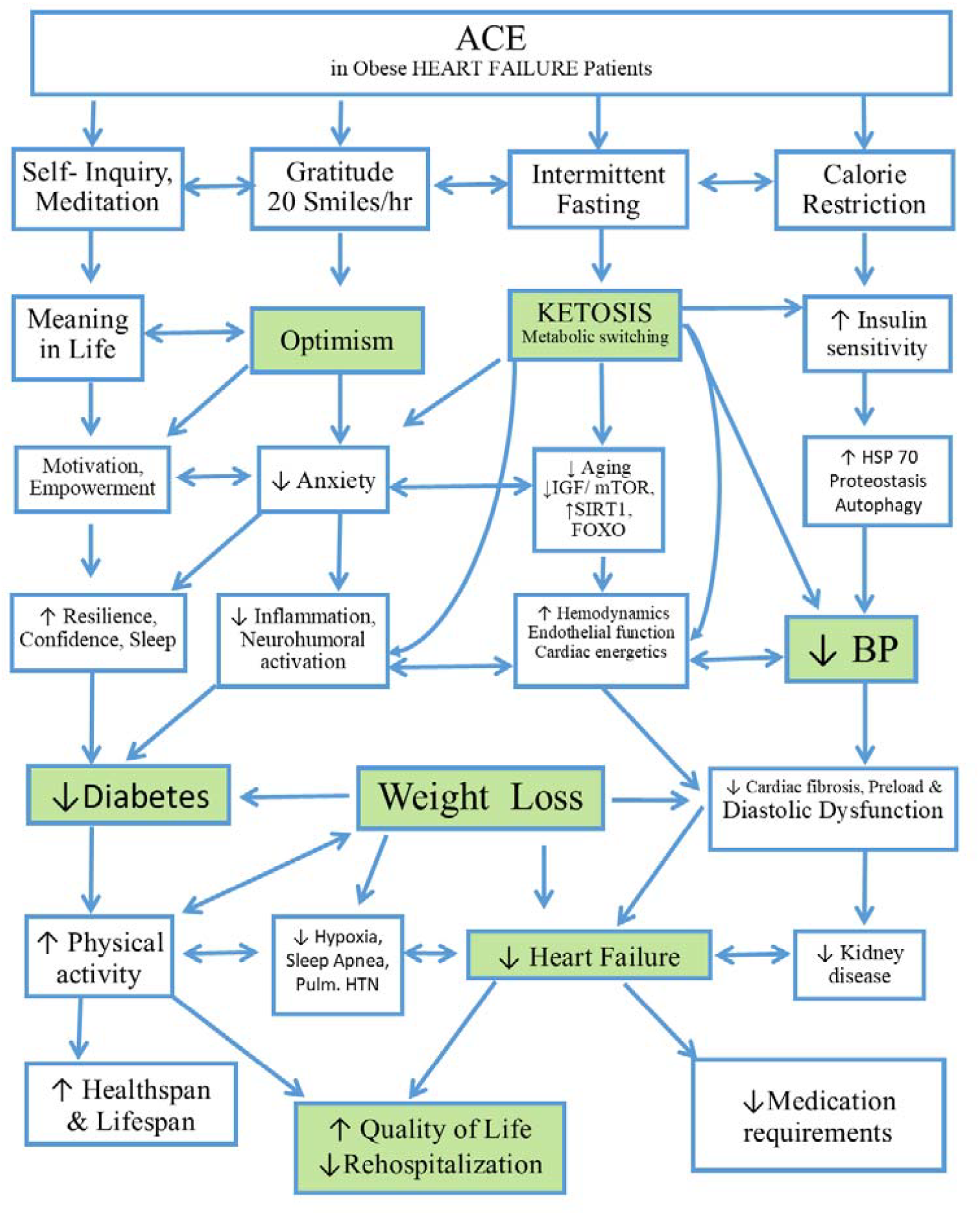
Advanced Cardiac Energetics (ACE) targets mental energy gains through self-inquiry and gratitude experience as well as mindful eating through fasting and calorie restriction with significant individual variability. There are several pathways that are likely implicated and synergetic accounting for the holistic empowerment achieved. The boxes highlighted in green are the mechanisms that routinely achieved in our pragmatic outcomes program.

## Data Availability

All data produced in the present work are contained in the manuscript

